# Genetic analysis of Indian sporadic young onset patient with Amyotrophic Lateral Sclerosis

**DOI:** 10.1101/2023.09.12.23295297

**Authors:** Saileyee Roychowdhury, Deepika Joshi, Vinay Kumar Singh, Mohammed Faruq, Parimal Das

## Abstract

**Background:** Amyotrophic lateral sclerosis (ALS) is an old onset devastating neurodegenerative disorder. Young-onset ALS cases especially sporadic ones who are between 25 and 45 years are rarely affected by the disease. Despite the identification of numerous candidate genes associated with ALS, the aetiology of the disease remains elusive due to extreme genetic and phenotypic variability. The advent of affordable whole exome sequencing has opened new avenues for unraveling the disease’s pathophysiology better.

**Methods and Results:** Our aim was to determine the genetic basis of an Indian-origin, young onset sporadic ALS patient with very rapid deterioration of the disease course without any cognitive decline who was screened for mutations in major ALS candidate genes by Whole exome sequencing. Variants detected were reconfirmed by Sanger Sequencing. The clinicopathological features were investigated and two heterozygous missense variants harboring each in *ANXA11,* in one of the four conserved C terminal domains (R452W) and another in the *SIGMAR1* (R208W) were thus identified respectively. Both of these variants were predicted to be damaging by pathogenicity prediction tools and various *In silico* methods.

**Conclusions:** Our study revealed two potentially pathogenic variants in two ALS candidate genes. The genetic makeup of ALS patients from India has been the subject of a few prior studies, but none of them examined *ANXA11* and *SIGMAR1* genes so far. These results establish the framework for additional research into the pathogenic processes behind these variations that result in sporadic ALS disease and further our understanding of the genetic makeup of Indian ALS patients.

## 1. Introduction

Amyotrophic lateral sclerosis is a late onset, devastating, multisystem brain neurodegenerative disorder, affecting the upper and lower motor neurons of the body. Phenotypically characterized by general weakness, muscle stiffness and spasticity, muscle atrophy, brisk reflexes, fasciculations, dysarthria, and problems in swallowing [1]. This rapid progressive disorder causes death from respiratory failure within 3-5 months of the disease onset [2], [3], [4]. Around 5-10% of cases are classed as familial ALS (fALS) cases because they have a positive family history, while the other 90-95% of cases with complex inheritance patterns are sporadic in nature (sALS) and lack a clear family history [5]. ALS has a global frequency/prevalence of 4.42 cases per 100,000 people and an annual incidence of around 1.59 cases per 100,000 people [6]. About 5-10% of ALS cases are presented with symptoms of frontotemporal dementia (FTD) and thereby falls into ALS-FTD spectrum of disease [7].

More than 30 major candidate genes have been identified to date to be linked to the disease onset [8]. The most frequently mutated are being *Sod1*, *C9orf72*, *FUS* and *TARDBP* [9]. Mutations in these genes affects major molecular pathways that lead to protein misfolding and their subsequent aggregation, impaired RNA processing, glutamate excitotoxicity, oxidative stress, mitochondrial dysfunction, and impaired axonal transport thus leading to motor neuron degeneration [10].

Recently in 2017, *ANXA11* has been included in the list of ALS causing candidate genes [11]. *ANXA11* which is ubiquitously expressed in different cell types encodes Annexin A11, which is a calcium-dependent phospholipid-binding protein [12], [13] with a molecular weight of 56 kDa and consisting of 504 amino acids [14]. It has an stabilising N terminal low complexity domain which is rich in Proline, Glycine, and tyrosine residues and are involved in liquid-liquid phase separation (LLPS) [15]. It has a binding site for a calcium-binding protein called calcyclin which is encoded by S100A6 and also for programmed cell death protein 6 encoded by PDCD6 respectively, whereas the C terminal region with 4 homologous Conserved Annexin domains of approximately 70 amino acid residues each [12], are responsible for phospholipid binding in a calcium-dependent manner and are also involved in Ca^2+^ signalling, cell division, apoptosis, and vesicle trafficking [11], [16]. Several studies indicate that Annexins play a function in autoimmune illnesses like sarcoidosis [17],[18] cardiovascular disease [19], and cancer [20]. Recent research has found that several *ANXA11* mutations associated with ALS increase aggregation propensity, resulting in disruption of intracellular Ca2+ homeostasis and RNA granule trafficking [21].

*SIGMAR1:* The endoplasmic reticulum (ER) chaperone protein known as the sig-1 receptor (Sig-1R or **σ_1_R)** was initially identified as the sigma opioid receptor by Martin *et al.* in 1976 [22], however it was later discovered to be a unique nonopioid receptor [23]. This integral transmembrane protein encoded by *SIGMAR1*, is made up of 223 amino acids. Though expressed ubiquitously, in the neurological system, *SIGMAR1* is highly expressed in the CNS especially in the motor neurons of the brain stem and spinal cord and subcellularly enriched in postsynaptic locations [24]. This gene is involved in the regulation of various molecular functions in various cell types, including the calcium signalling, mitochondrial function, autophagy, ion channel function regulation, lipid transport and prevention of accumulation misfolded proteins [25]. Functionally **σ_1_R** modulates calcium signalling via inositol 1,3,5-triphosphate receptors (IP3 receptors) [26] and binds to a variety of ligands such as neurosteroids, stimulants, and detrobenzomorphans.

During Endoplasmic Reticulum (ER) stress, σ_1_R translocate from mitochondria-associated ER membrane (MAM), which serves as the interface between the ER and mitochondria to the plasma membrane [27]. While σ_1_R dysfunction and its related mutations have been linked to a number of neurological diseases like Alzheimer’s disease, Parkinson’s disease, Huntington’s disease, Amyotrophic lateral sclerosis, Distal hereditary motor neuropathies, and schizophrenia, σ_1_R activation using ligands induces potent neuroprotective effects, improves neuronal survival, and restores neural plasticity to halt disease progression [27]–[29].

The presence of *ANXA11* and *SIGMAR1* variants have been screened in a number of cohorts from different geographic regions. While prior research has explored the genetic makeup of ALS patients in India, our study stands out by its examination of the *ANXA11* and *SIGMAR1* genes along with other ALS candidate genes, contributing to a deeper understanding of sporadic ALS and the genetic diversity within the Indian ALS patient population [30]–[32].

Young onset ALS cases have a distinct genotype and phenotypic characteristics than their older counterparts. Recent research has indicated that young-onset ALS may be more genetically predisposed than other ALS cases. *SPG11*, *ALS2*, *SETX* and *FUS* variants are the ones that are frequently altered in these cases. However, in these cases the likelihood that other most frequently occurring genes such as *SOD1, TARDBP*, *C9orf72* are relatively less [33],[34],[28]. Consistent with other studies, young onset ALS cases has a spinal onset male preponderance [29], [33]. However, to fully comprehend the genetic makeup of the patients with young-onset ALS, further research is needed in future.

In this present study, we describe variations in *ANXA11* and *SIGMAR1* in a young onset ALS patient and demonstrate the impact of this mutation on the respective genes’ normal function by *In silico* studies.

## 2. Materials and Method

### 2.1. Sample selection

A young male patient in early 20s (21-25 years) with a disease course of 14 months from Northern parts of India with no reported family history of neurological disorder, presented with abnormal movement of left lower limb followed by right lower limb, spasticity, speech difficulty along with atrophic hand and legs, abnormal posturing of his left hand fingers mainly, index, middle and ring fingers, weakness in the limbs and restricted movement was admitted to the neurology ward of Institute of Medical Sciences, Banaras Hindu University, India a tertiary care academic hospital in Uttar Pradesh. Initial symptoms, time of disease duration were recorded as reported by the patient and his relatives after obtaining informed written consent. The patient did not have any complaints suggesting dementia.

### 2.2. Clinical assessment

The proband and his mother provided information on the family history. The proband underwent electromyography (EMG) testing, nerve conduction velocity studies (NCV) of both bilateral upper and lower limbs, and magnetic resonance imaging (MRI) of his head and cervical cord. The proband also had additional laboratory testing, such as an ECG, full blood count, Ca2+ and P+, thyroid profile, vitamin B12, serum homocysteine. The neuropathological features were in accordance with the ALS El Escorial scale and was classified as a Definite ALS case as confirmed by EMG and NCV by a senior neurologist. The severity of the disease was calculated using the ALS Functional Rating Scale-Revised (ALSFRS-R). The study was conducted in accordance with the Declaration of Helsinki and the was approved by the Institutional Ethics Committee, Banaras Hindu University.

### 2.3. DNA isolation : Genetic analysis

Under aseptic conditions, 5 ml of peripheral blood was collected using a heparinized needle and stored in an anticoagulant (K2 EDTA) precoated collecting tube. Genomic DNA was extracted using salting out method. The quality and quantity of the DNA in the isolated sample was evaluated using Nanodrop 2000 (Thermo Fisher Scientific Inc., USA) while the integrity and quality of the sample was checked by running it on 0.8% agarose gel.

The presence of pathogenic G4C2 hexanucleotide repeat expansions in *C9orf72* was tested by Repeat-primed PCR assay along with fragment analysis [35] as well as trinucleotide CAG repeat sizes in *ATXN*-1 and *ATXN*-2 were also tested by PCR amplification by Fluorescently labelled primers [36] in the patient sample.

### 2.4. Next Generation Sequencing

Whole exome sequencing including all coding exons with extended UTR regions, splice sites, and extended intron regions (30bp) immediately adjacent to the alternatively spliced exons was performed by Illumina Hi-seq or Nova Seq 6000 sequencer using the Agilent V6 + UTR reagent kit (Agilent Technologies, Santa Clara, CA) with 150X2 paired end read cycles. The quality check of raw reads obtained from the next generation sequencing platform was checked using FastQC (v0.11.5) software (https://www.bioinformatics.babraham.ac.uk/projects/fastqc/). It was followed by the removal of the adapter using FastX-ToolKit (v0.0.13) (http://hannonlab.cshl.edu/fastx_toolkit/). Mapping of Paired-end raw reads to the Human Reference Genome (UCSC Genome Browser build hg19) was done using Bowtie2 software (v2.2.6) (http://bowtie-bio.sourceforge.net/bowtie2/index.shtml) followed by the variant calling and annotation, performed by using GATK (v3.4.46) (https://gatk.broadinstitute.org/hc/en-us) and SnpEff (v4.3) (http://snpeff.sourceforge.net/SnpEff_manual.html) or Annovar (v24/10/2019) (https://doc-openbio.readthedocs.io/projects/annovar/en/latest/), respectively. Variants were classified and interpreted according to American College of Medical Genetics and Genomics (ACMG) guidelines.

For identifying the genes responsible for disease occurrence, variants were further selected from the annotated VCF file following these criteria: read depth of at least 30×, Minor allele frequency (MAF) < 1% in several population databases like Genome Aggregation Database (GnomAD) (https://gnomad.broadinstitute.org/) [37], ExAC (http://exac.broadinstitute.org/), 1000genome database (https://www.internationalgenome.org/), Human gene mutation database (HGMD) (https://www.hgmd.cf.ac.uk), database of single nucleotide polymorphism (dbSNP) (http://www.ncbi.nlm.nih.gov/projects/SNP), Exome variant server (https://evs.gs.washington.edu) . Pathogenicity prediction: The plausible effects of the identified variants found were determined initially through available online *In silico* tools such as SIFT [38], PolyPhen-2 [39], Mutation taster [40], SNPs&GO [41] and PROVEAN [42] respectively. Variants that were non synonymous in nature and classified as pathogenic, likely pathogenic and variants of uncertain significance (VUS), were only considered for the study.

### 2.5. Sanger sequencing

Primers were designed for exon containing specific mutations for Polymerase chain reaction (Thermal cycler, ABI). After PCR amplification the amplified product was purified by Exonuclease and Shrimp Alkaline Phosphatase (USB Affymetrix, USA) and directly sequenced on an ABI 3130 automated sequencer (Applied Biosystems, CA, USA) for reconfirmation of the proband’s DNA sequence variants found through NGS. Sequencing reads were then compared against NCBI BLAST.

### 2.6. Evolutionary conservation analysis

For the ANXA11 and SIGMAR1 proteins, the evolutionary conservation of amnio acid residues was evaluated using the CONSURF algorithm. The homology search algorithm used was HMMER in 1 iteration with cut off of 0.0001 for the E-value. The protein database of choice was UNIPROT. For homologs, 95% and 35% were chosen as the maximum and minimum identities, respectively. The Multiple Sequence Alignment (MSA) was built using CLUSTALW as the alignment method. The scores were calculated according to Bayesian method.

### 2.7. Secondary structure prediction

For protein structure prediction, the PSIPRED programme (http://bioinf.cs.ucl.ac.uk/psipred/) was used which combines PSIPRED, GenTHREADER, and MEMSAT2 techniques. Two feed-forward neural networks are used in this prediction method to analyse the results of PSI-BLAST [43]. While the second network forecasts interactions between proteins and ligands, the first network detects residues that are crucial for defining a ligand’s position [43].

### 2.8. Homology modelling prediction

Since h*ANXA11* did not have a crystal structure, homology modelling using computational method was used to determine its three-dimensional (3D) structure along with R452W variant, using the I-TASSER [44], RaptorX [45] and LOMET programs. Through adjusting the protein sequence and interaction partners, a total of 5 models were produced. On the basis of the R-factor and Ramachandran plot, the model that best fit the experimental data was selected. In case of Sigmar1 variant its wild type structure was retrieved from PDB. Then 3D structure of R208W variant was predicted through homology modelling using I-Tasser. Structure refinement of the predicted model was done using ModRefiner [46]. SAVES v6.0 toolkit (https://saves.mbi.ucla.edu/) [47] comprising of 5 tools namely ERRAT, VERIFY3D, PROVE, PROCHECK and WHATCHECK was used to further validate the predicted models. This toolkit predicts several stereochemical properties of the protein structure by evaluating the Ramachandran plot or by using PROCHECK to analyze the geometry of the entire model as well as the geometry of each individual residue. In order to compare the 3D structural differences between the wild type and the variant type, the models were then visualized and superimposed using Biovia Discovery Studio Visualizer.

### 2.9. Molecular dynamics simulation studies

In order to identify the pathogenicity of the variants found, Molecular dynamics (MD) studies were performed on the wild type and the mutated type of *ANXA11* and *SIGMAR1* respectively. The models of wild type and its variant of both *ANXA11* and *SIGMAR1* were submitted to online WebGRO Macromolecular Simulations server (https://simlab.uams.edu/). The models were simulated for 100 ns by implying GROMOS96 43a1 forcefield and the protein fragment was solvated in SPS water molecules in a triclinic box and sodium chloride, neutralized and 0.15M salt of NaCl was introduced. Using the steepest descent method for every 5000 steps, the systems underwent energy minimization. The NVT/NPT equilibration was carried out at 300 K and 1 bar of pressure. The MD integrator used was Leap-frog with simulation time of 100ns and the number of frames per simulation were fixed at 1000 [48]. Main parameters such as RMSD (Root Mean Square Deviation), RMSF (Root Mean Square Fluctuation), were evaluated for complexes stability.

### 2.10. Prediction of change in stability due to variation in protein sequence

I-Mutant 2.0 (https://folding.biofold.org/i-mutant/i-mutant2.0.html), a support vector machine tool, was used to predict how the protein’s stability would change after a mutation. The I-Mutant 2.0 server, which automatically predicts changes in protein stability caused by single point mutations, was developed and tested using the most comprehensive thermodynamic experimental data library of free energy changes in protein stability due to mutation, ProTherm. I Mutant 2.0 can be used as a regression estimator by predicting the change in Gibbs-free energy (G) due to a point mutation, as well as a classifier for determining the direction of the change in protein stability [49].

## 3. RESULTS

### 3.1. Clinical assessment

The young male patient with complaints of weakness of all four limbs and difficulty in speech for last 14 months was admitted. The patient had the limb as the first place of disease onset. The patient, who was a nonsmoker and non alcoholic was apparently alright fourteen months back from the time of admission when he first developed weakness of left lower limb in the form of slipping of his slippers from his foot with awareness but could perform his daily activities. He had insidious onset of symptoms which slowly became progressive after few months. He gradually had weakness of left upper limb with difficulty in holding small objects. The symptoms gradually progressed to right lower limb and then to right upper limb whereby he performing any of his daily activities became challenging. It was progressed further to bulbar regions with speech difficulty and swallowing problems. The patient also complained of increased stiffness of his upper and lower limbs. There was a marked atrophy of the muscles of his hands and legs. There was no history of any numbness or tingling sensation or any history of burning sensation over the body.

On history of clinical examination, the patient was found to have gradually progressive spastic asymmetric quadriparesis, upper motor neuron (UMN) type with UMN type speech with pseudobulbar effect with visible fasciculations over tongue and upper limbs without any sensory or cranial involvement with normal bowel and bladder. NCV and EMG studies confirmed the case to be a definite young onset ALS (UMN and LMN quadriparesis). ALSFR score measured was 21 out of 40.

### 3.2. Genetic results

Two heterozygous missense variation, one located in exon 16 of *ANXA11* (c.1354>T) altering the amino acid at 452 position from Arginine to Tryptophan (p.R452W) and the other missense variant (c.622C>T, p.R208W, rs11559048) altering the amnio acid at 208 position from Arginine to Tryptophan in exon 4 of the *SIGMAR1* respectively were identified in the young onset ALS patient sample. In both the cases, a hydrophilic and basic amnio acid, Arginine got converted into a hydrophobic and aliphatic amino acid, Tryptophan. The *ANXA11* variant (p.R452W) was present in one of the conserved homologous domains in the C terminal region. It has been found that patients with variations in the conserved ANX domain suffer from more classical phenotypes than the bulbar onset [50], which was also seen in our patient. No other variants were found in other ALS candidate genes. The Minor Allele Frequency (MAF) of *ANXA11* p.R452W is <0.01 while the p.R208W in *SIGMAR1* has a MAF of 0.007 (global) and 0.019 (South Asian) according to ExAC database respectively. The Phred-scaled Combined Annotation Dependent Depletion (CADD) score for both the variants were > 30. Both the variations were cross confirmed by Sanger Sequencing (Fig: 2a, 2b).

**Fig: 1.**
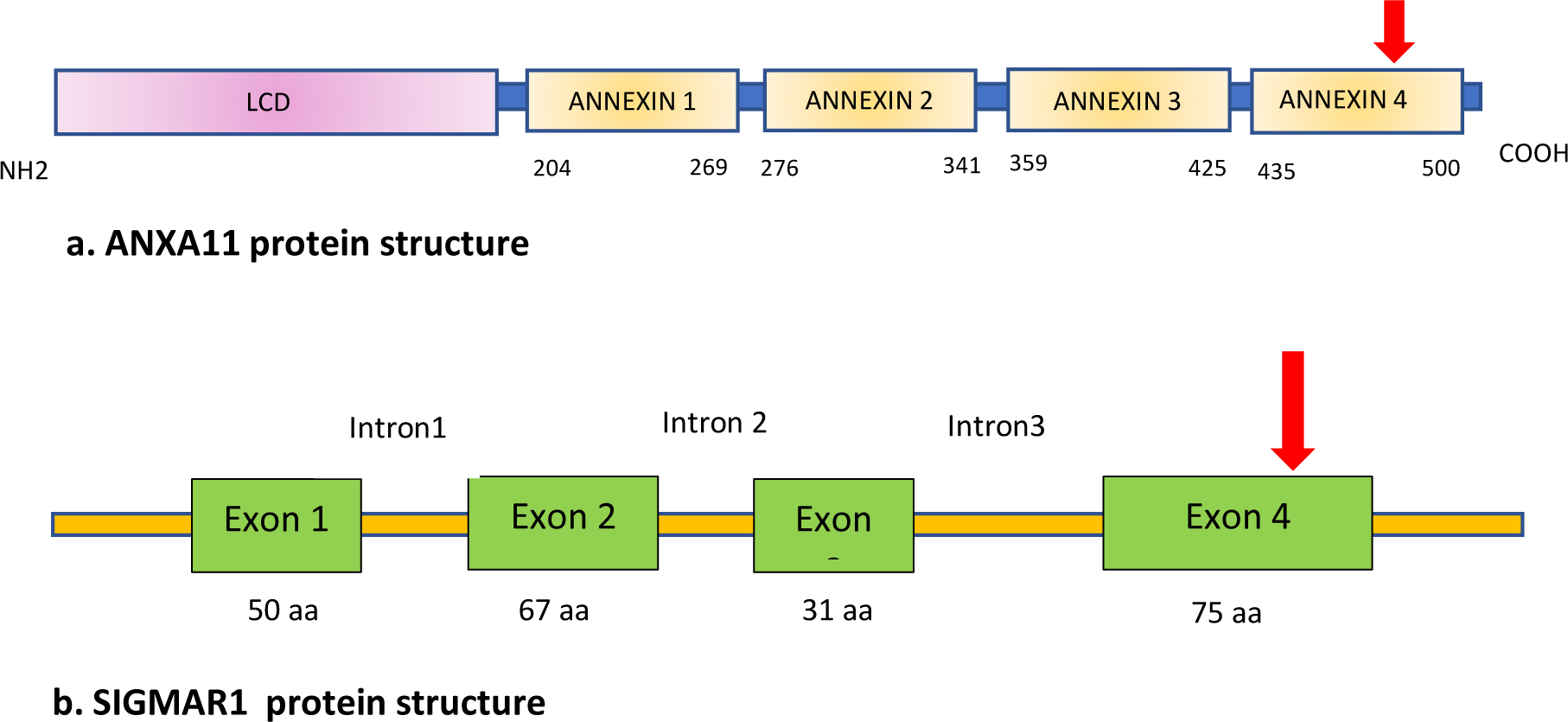
Graphical representation of **(a)** the domains present in the ANXA11 protein. LCD represents low complexity domain which is a binding site for another calcium-binding protein, Calcyclin.in the extended N - terminal region. Annexin 1 Annexin 2, Annexin 3, Annexin 4 represents conserved homologous domains associated with phospholipid binding via calcium regulation in the in the C terminal region. The Red arrow indicate the position of p.R452W mutation **(b)** the exons with the number of amino acids respectively and introns present in the SIGMAR1 protein. The red arrow indicates the position of p.R208W mutation

**Fig: 2.**
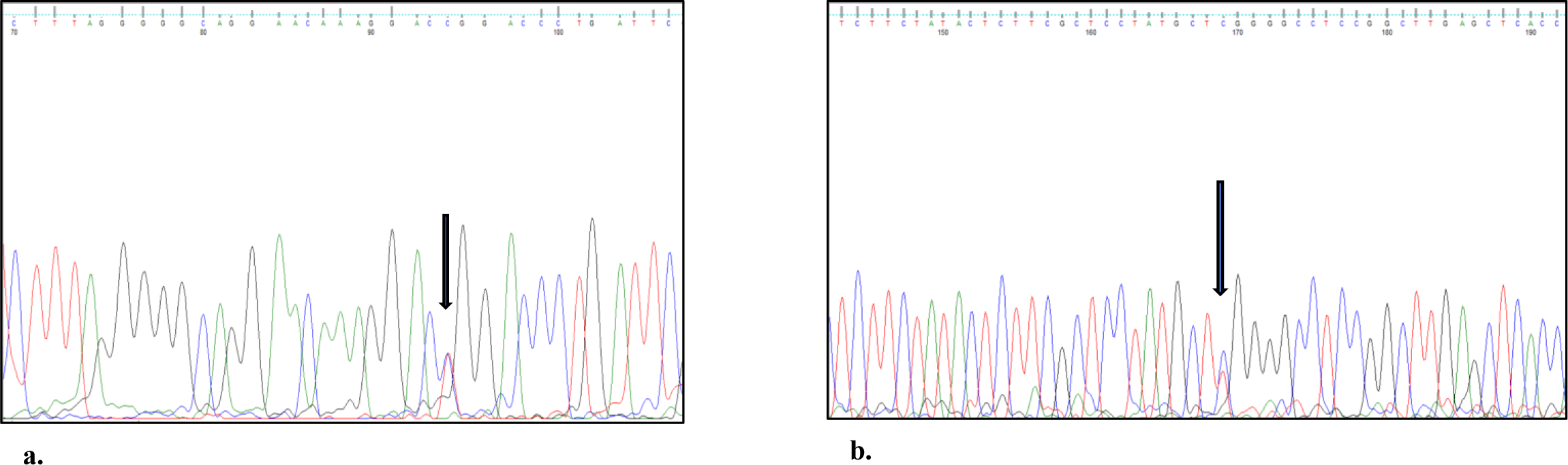
Chromatograms showing changes in **(a)** *ANXA11* c.1354C>T change (p.R452W) and **(b)** *SIGMAR1* c.622C>T (p.R208W)

### 3.3. Pathogenicity prediction

Several different *In silico* pathogenicity prediction algorithms such as Mutation Taster, SIFT [38], Polyphen and PROVEAN that were used to predict the functional and structural effects of these variants at the protein level, predicted both R452W and R208W to be deleterious in nature as shown in Table 1.

**Table 1:** Pathogenicity prediction of ANXA11 R452W and SIGMAR1 R208W variants by different *In silico* tools.

| Pathogenic Prediction tools | *ANXA11* p.R452W | *SIGMAR1* p.R208W |
| --- | --- | --- |
| SIFT | Deleterious | Deleterious |
| PROVEAN | Deleterious | Deleterious |
| PolyPhen | Deleterious | Deleterious |
| Mutation Taster | Deleterious | Deleterious |
| LRT | Neutral | Deleterious |
| SNPs&GO | Deleterious | Deleterious |

### 3.4. Conservation analysis

Consurf analysis showed that though the R452W variant in *ANXA11* is present in one of the four conserved domains in the C terminal region, however the amino acid Arginine (R) with a conservation score of 2 out of 10 is predicted to be variable in nature (Fig: 3a). The R208W variant had a score of 5 out of 10 denoting that it is averagely conserved across species. (Fig: 3b), therefore any mutation at that position might hamper its function as well.

**Fig: 3a.**
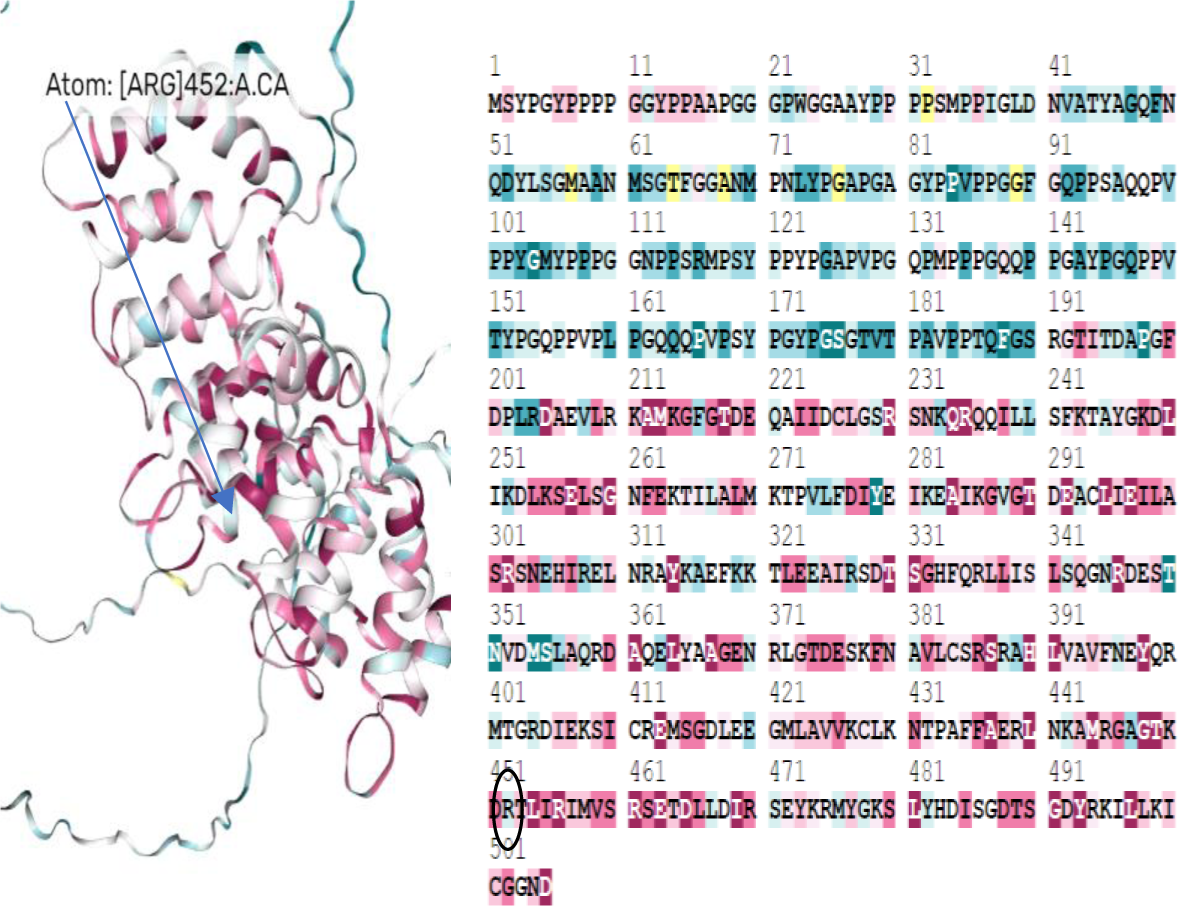
Conservational analysis of ANXA11 wild. Conservation score= 2

**Fig: 3b.**
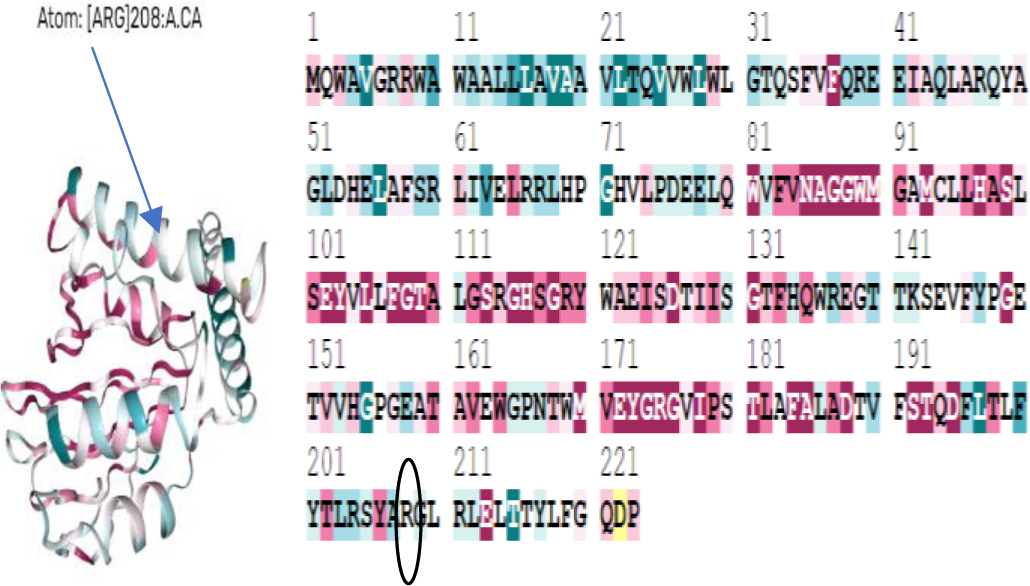
Conservational analysis of SIGMAR1 wild. Conservation score= 5

### 3.5. Bioinformatic analysis

Secondary structure prediction: Analysis of the secondary structure by PsiPred showed significant sites of alterations both at upstream and downstream positions from the site of variation in the mutated R452W *ANXA11*. These changes included shortening of a helical structure (431-432 aa and 451-457 aa)) and increase in the strand structure (458-460) (Fig: 4) whereas a slight change was found in case of mutated R208W *SIGMAR1* variant. (Fig: 5)

**Fig: 4.**
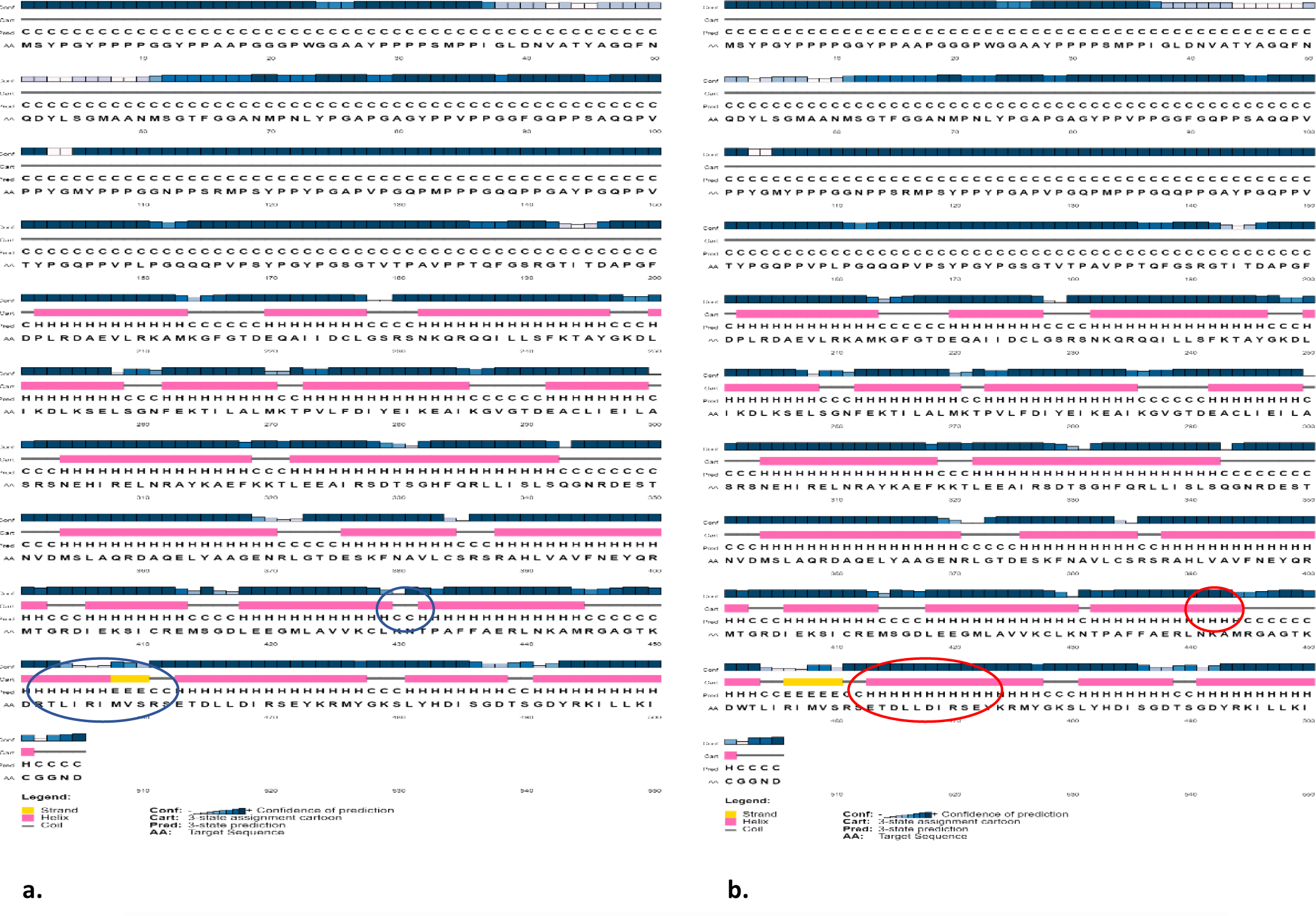
**(a)** Site of alterations shown in circle in the predicted secondary structure of a. *ANXA11* wild type **(b)** *ANX11* R452W type by PSIPRED

**Fig: 5.**
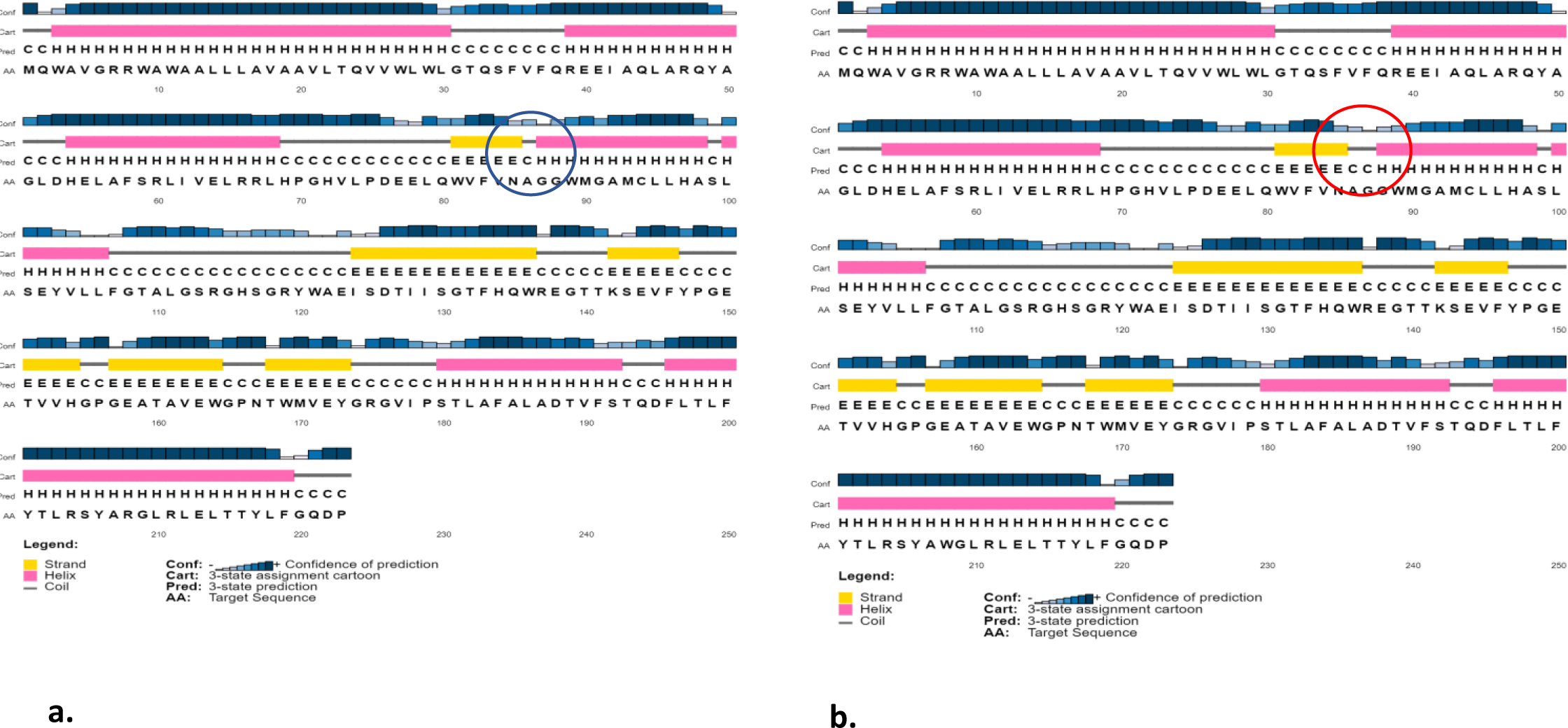
**(a)** Predicted secondary structure of a. *SIGMAR1* wild type **(b)** *SIGMAR1* R208W type by PSIPRED

### 3.6. Prediction of Protein stability

Using the software, I-Mutant 2.0, it was found that there was a decrease in stability of the amino acid substitution from Arginine to Tryptophan at 452 position in *ANXA11* gene (ΔΔG= -0.77) and 208 position in SIGMAR1 gene (ΔΔG= -0.37) respectively at pH 7.0 and temperature 25C.

### 3.7. Physiochemical properties

The characteristics of individual amino acid that constitute a protein are greatly influenced by their physiochemical properties. Since, every change in an amino acid at a given position has the potential to affect the physiochemical properties of that respective protein, our next goal was to investigate how the R452W and R208W mutations affected the overall physiochemical characteristics of wild *ANXA11* and *SIGMAR1* respectively. Therefore, Protscale software was used which predicted the mutant p. R452W to have lower hydropathicity (Fig: 6a and 6b), increased hydrophobicity (Fig: 6c and 6d), decrease in polarity (Fig: 6e and 6f) and slight increase in alpha helix structure (Fig: 6G and 6H) when compared to *ANXA11* wild type within the region of amino acid substitution whereas p.R208W was found to have increased hydropathicity and hydrophobicity, lower polarity and slight increase in the alpha helix structure when compared to the wild type *SIGMAR1* (Fig: 7a-7f). In spite of having one amino acid substitution in the protein structure, the overall molecular weight increased in both p.R452W and p.R208W variant types when compared to their wild types. Instability index showed that both *SIGMAR1* wild type and p.R208W variant are stable in nature (instability index<40) while both wild type *ANXA11* and its variant p.R452W are instable in nature (instability index >55). Aliphatic index (AI) refers to the proportion of a protein’s volume that is occupied by its aliphatic side chains which in return influences the thermal stability of proteins. Proteins are more thermally stable and tend to have an increased hydrophobicity when their aliphatic index is high. Though the aliphatic index was same in both the cases when compared to their wild counterparts however the very high aliphatic index of *SIGMAR1* and its mutated type p.R208W indicates that they might be thermally stable over a wide range of temperature. There was a slight increase in the Grand average of hydropathicity (GRAVY) value which measures the sum of hydropathy values for all amino acids divided by the length of the sequence, in p.R452W with respect to *ANXA11* wild type, though both of them are slightly hydrophilic intrinsically in nature with negative GRAVY value indicating that the surface accessibility of the mutated protein might be affected, whereas *SIGMAR1* and R208W are slightly hydrophobic in nature. The isoelectric point (Theoretical pI) of R452W (6.91) indicated that it has become more acidic from basic than its wild type with theoretical pI measuring 7.53 while R208W of *SIGMAR1* has become slightly acidic when compared to its wild type (Table 2.).

**Fig: 6.**
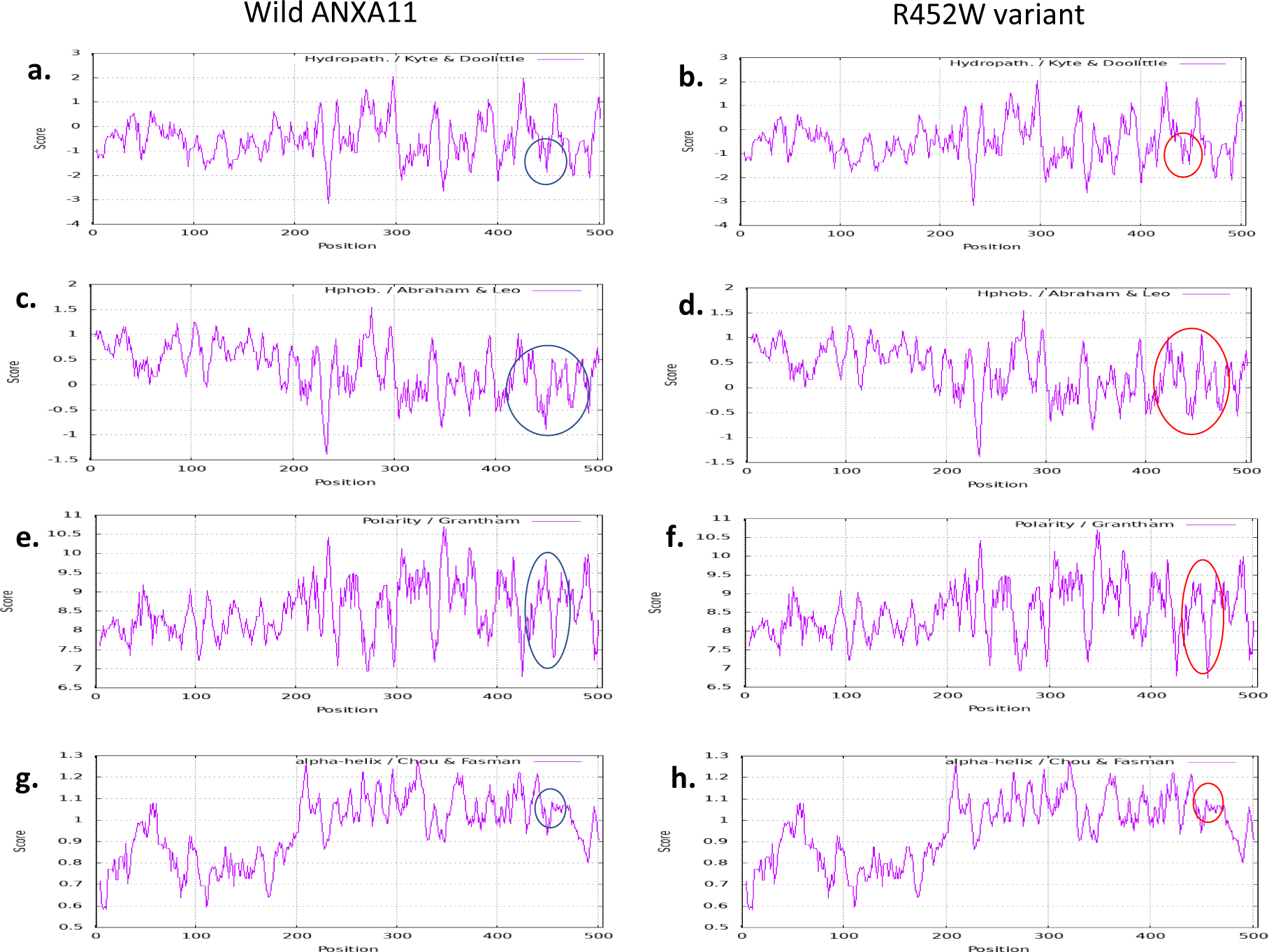
Physiochemical properties of *ANXA11* wild vs R452W variant. **(a,b)** denotes lower hydropathicity of R452W compared to its wild type **(c,d)** denotes increased hydrophobicity of R452W compared to its wild type **(e,f)** denotes lower polarity of R452W compared to its wild type **(g,h)** denotes slight increase in alpha helix of R452W compared to its wild type. The x-axis represents the position of the amino acids while the y-axis represents the score in a default window size of 9

**Fig: 7.**
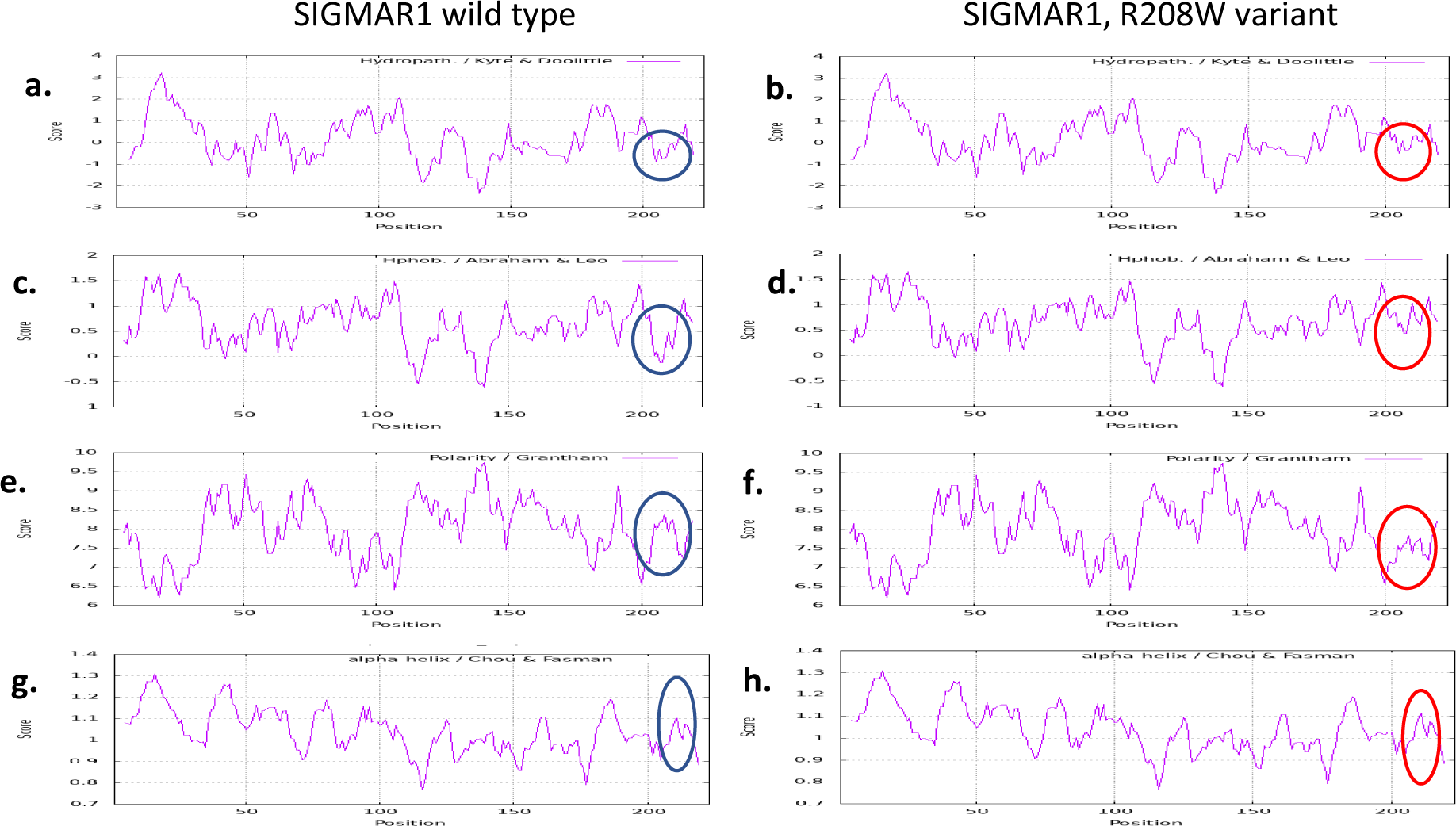
Physiochemical properties of *SIGMAR1* wild vs R208W variant. **(a,b)** denotes increased hydropathicity of R208W compared to its wild type. **(c,d)** denotes increased hydrophobicity of R208W compared to its wild type. **(e,f)** denotes lower polarity of R208W compared to its wild type. **(g,h)** denotes slight increase in alpha helix of R208W compared to its wild type. The x-axis represents the position of the amino acids while the y-axis represents the score in a default window size of 9

**Table 2:** Differences in physiochemical properties of wild type and its variant of *ANXA11* and *SIGMAR1*.

| Physiochemical Properties | *ANXA11* wild | *ANXA11* R452W | *SIGMAR1* wild | *SIGMAR1* R208W |
| --- | --- | --- | --- | --- |
| Molecular weight | 54389.71 | 54419.74 | 25127.69 | 25157.72 |
| Insatbility index | 58.66 | 58.51 | 34.02 | 33.94 |
| Aliphatic index | 64.97 | 64.97 | 94.48 | 94.48 |
| Grand average of hydropathicity (GRAVY) | - 0.521 | -0.514 | 0.134 | 0.150 |
| Theoretical pI | 7.53 | 6.91 | 5.61 | 5.45 |

### 3.8. Homology modelling

The three-dimensional protein structure of both the wild type *ANXA11* and *SIGMAR1* and its variant p.R452W and p.R208W were modelled and validated. PROCHECK Ramachandran plot analysis of both the wild and mutated type of *ANXA11* and *SIGMAR1* showed that most of the amino acids are in favoured and allowed regions (Fig: 8) . Moreover, the superimposed structure of p.R452W and p.R208W with their wild types further showed changes in three-dimensional structure due to the variation (Fig: 9).

**Fig: 8.**
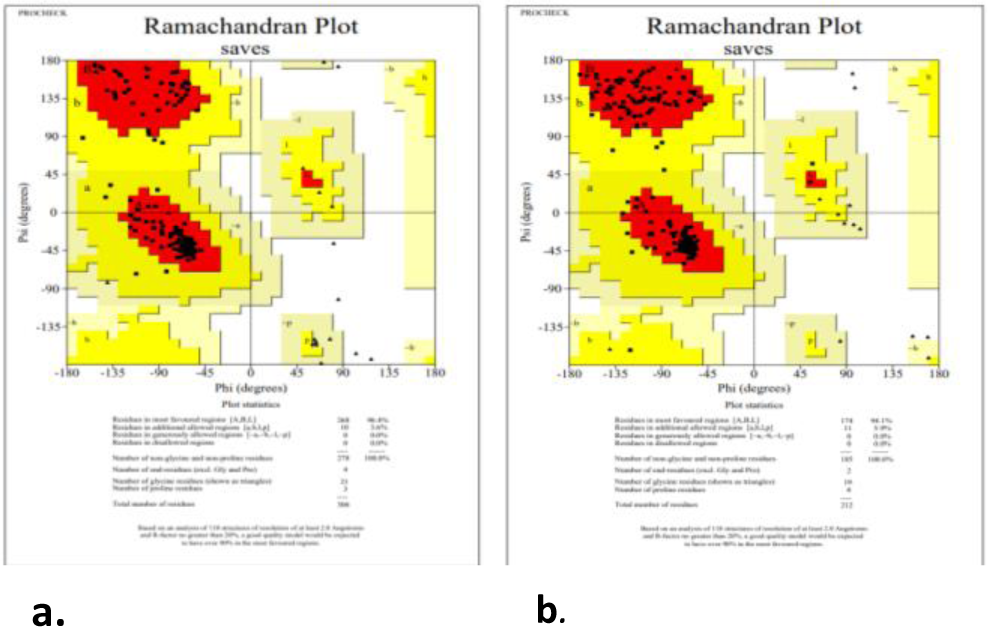
**(a)** Ramachandran plot of (a) *ANXA11* R452W model and **(b)** *SIGMAR1* R208W model

**Fig: 9.**
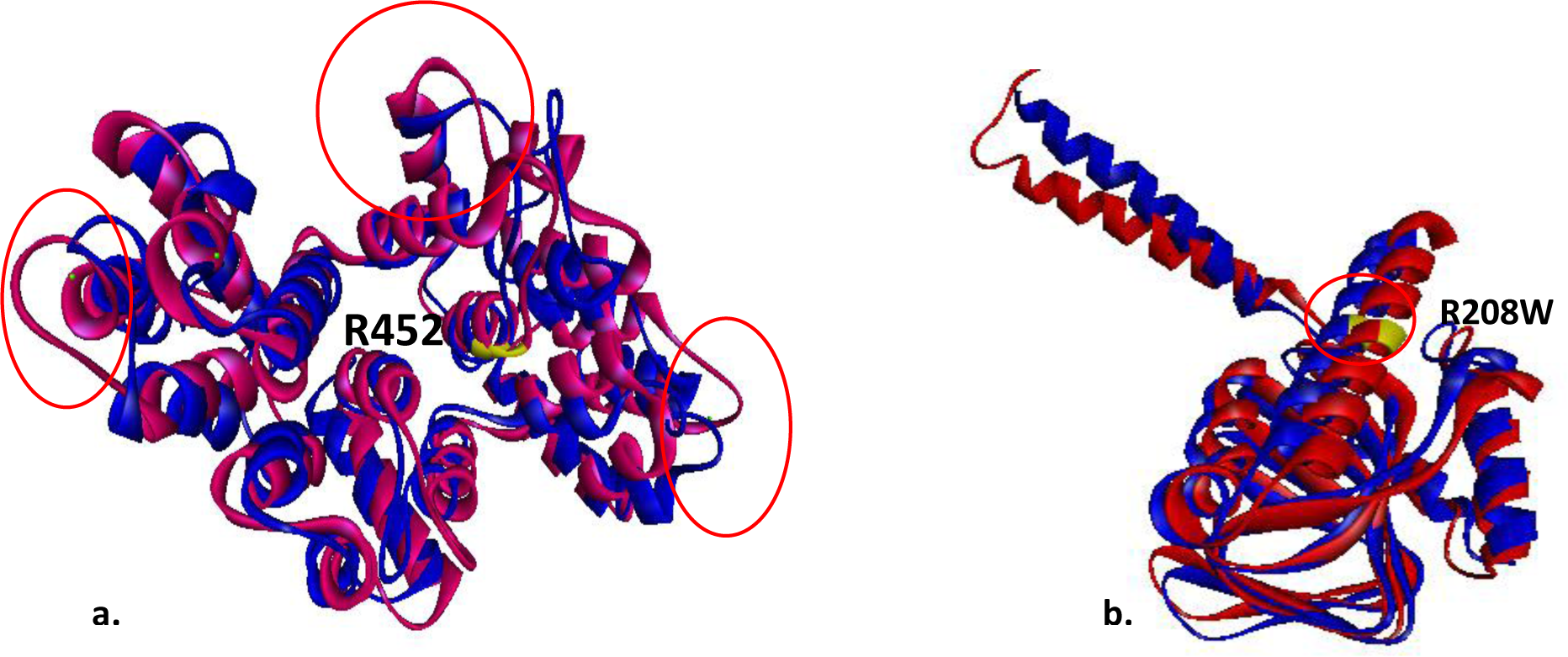
Superimposed structure of wild and variant type of **(a)** *ANXA11* and **(b)** *SIGMAR.* Blue denotes the wild type while Red denotes the mutated variant

**Fig: 10.**
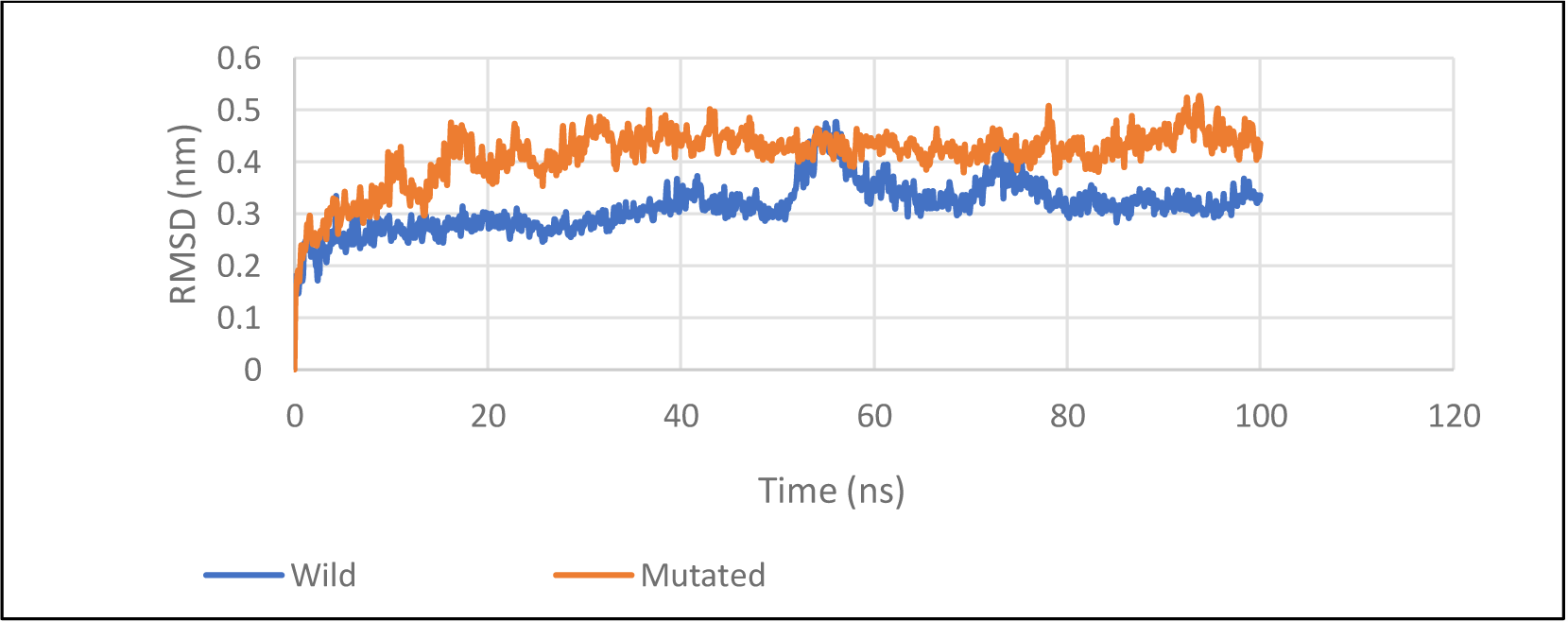
RMSD analysis of wild type *ANXA11* and its variant (R452W) over time (ns)

**Fig: 11.**
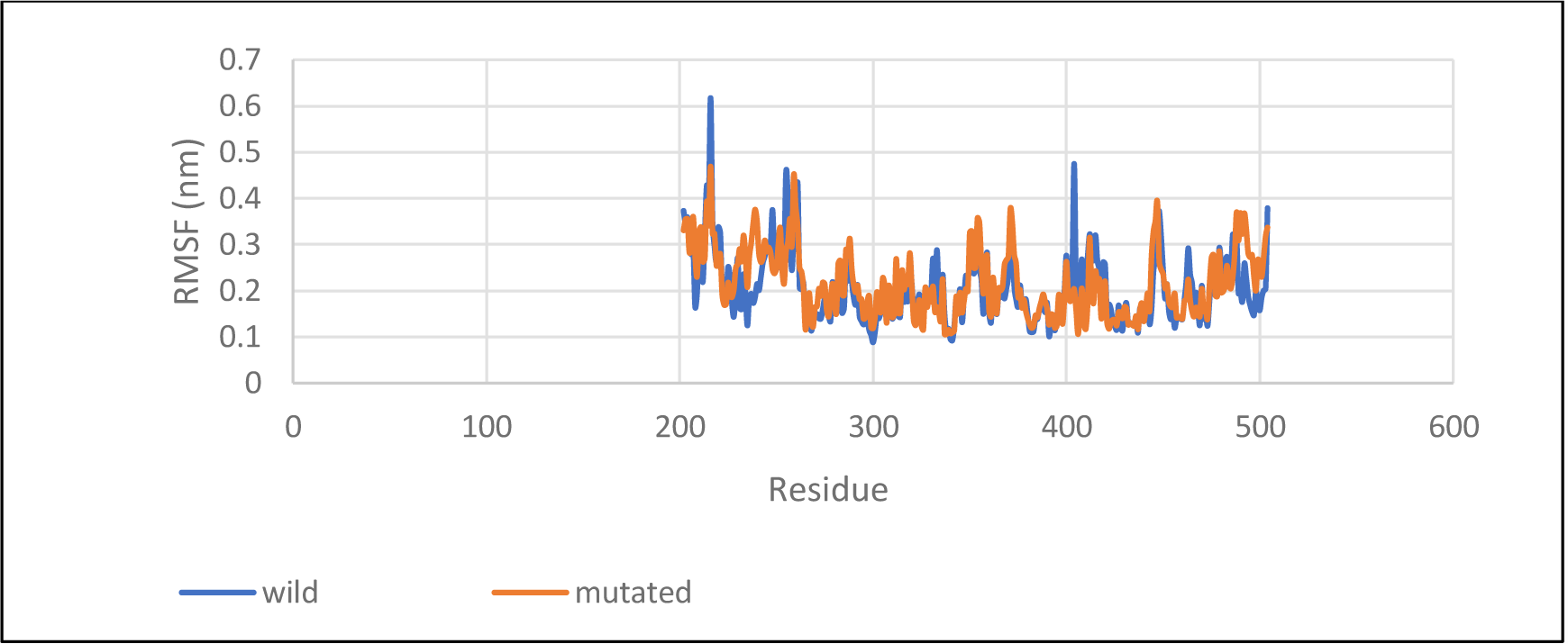
RMSF analysis of wild type *ANXA11* and its variant (R452W)

**Fig: 12.**
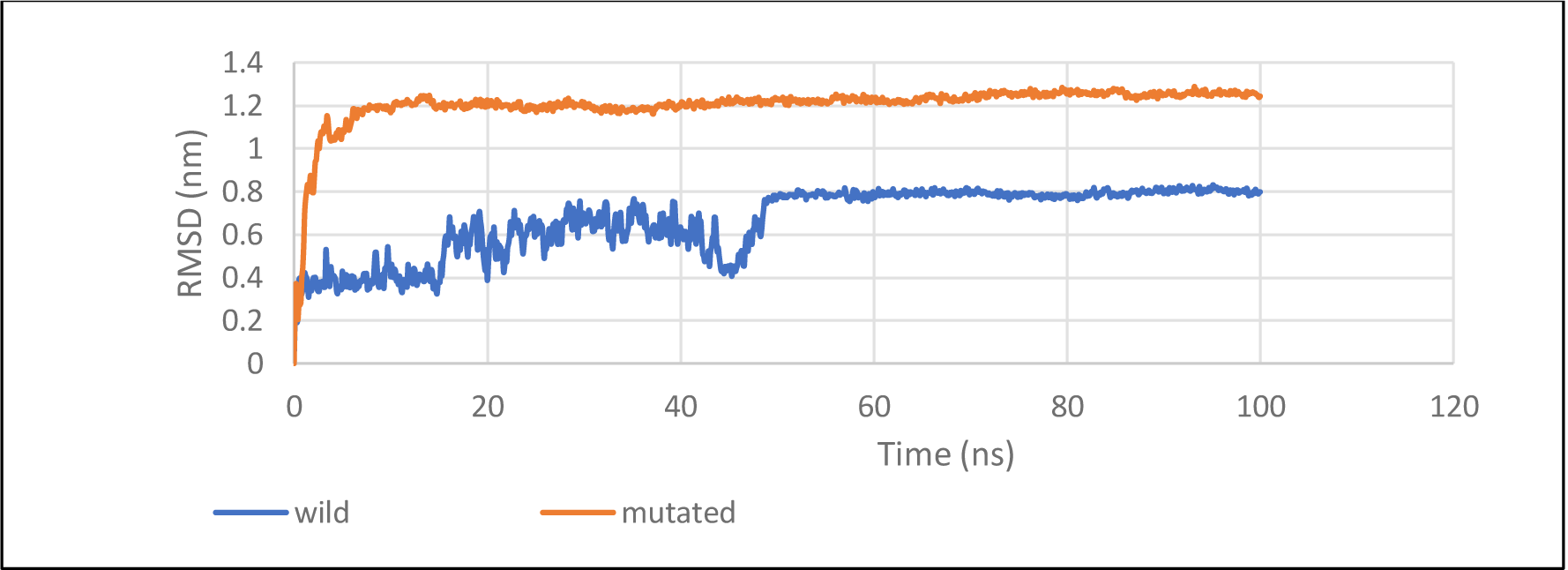
RMSD analysis of wild type *SIGMAR1* and its variant (R208W) over time (ns)

**Fig: 13.**
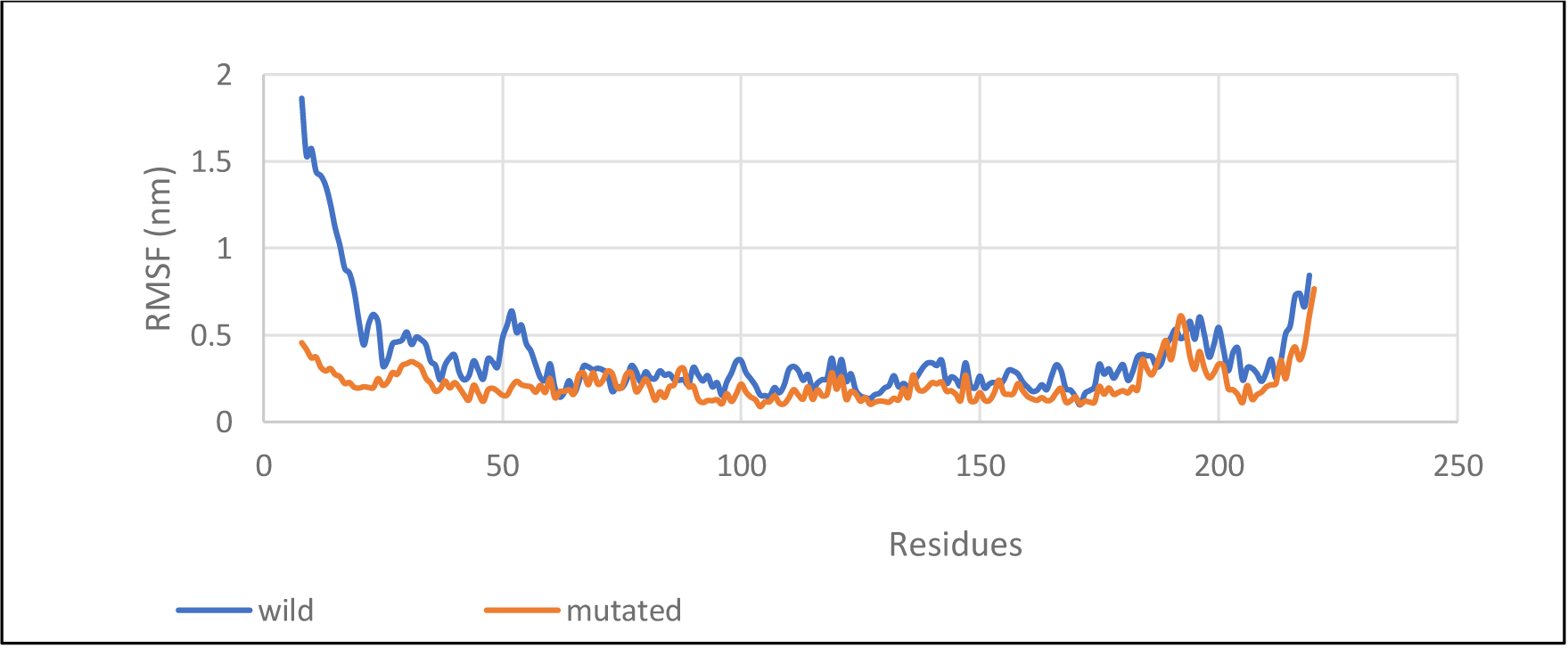
RMSF analysis of wild type *SIGMAR1* and its variant (R208W)

### 3.9. Molecular Dynamics simulation result

The data from the modelled superimposed structure were further supported by molecular dynamics simulation studies. Root mean square deviation is a commonly used indicator of structural separation between coordinates. It calculates the typical separation between a collection of atoms (backbone atoms of a protein). The RMSD value of two sets of protein structures (wild type and variant type) thereby indicates how much the protein shape has changed at two time points from the trajectory. Root mean square Fluctuation measures a particle’s average deviation from a reference point over time. Thus, RMSF analyses the structural elements that deviate most from their mean (or least).

Between the aligned structures of the wild type and its variation in *ANXA11* and *SIGMAR1* respectively, the RMSD and RMSF values demonstrate a considerable difference as depicted in Fig (10-13).

## 4. DISCUSSION

The high degree of clinical and genetic variability among different ALS patients along with the absence of any biological markers makes the diagnosis extremely difficult*. SOD1 FUS*, *TDP43* and *C9orf72* are the most frequently implicated genes in around two thirds of familial ALS cases. Since the majority of these genes are often inherited in an autosomal dominant manner, sporadic occurrence is extremely unlikely to be caused by them.

However, genes such as *SPG11*, *ALS2*, *SETX* and *FUS* have been linked to the rare forms of young onset ALS cases [33], [34], [51]. *ANXA11* and *SIGMAR1* have been recently identified as ALS candidate gene and several disease-causing mutations have been identified in these genes so far [24], [52]. Previous studies have delved in to the genetic composition of ALS patients in India but mutations in *ANXA11* and *SIGMAR1* has not been screened in Indian population so far. Our study distinguishes itself by screening not only previously reported candidate genes in India but also screening *ANXA11* and *SIGMAR1along with* other major ALS candidate genes

In the present study, Next generation sequencing identified a missense, non-synonymous variant in one of the conserved Annexin domain located in exon 16 of *ANXA11* along with another nonsynonymous variant in *SIGMAR1* in an aggressive young onset ALS patient with negative cognitive impairment. No other variants were found in other ALS candidate genes including pathogenic and intermediate repeat expansions in *C9orf72*, *ATXN1* and *ATXN2,* respectively.

Previous studies have found that mutations in *ANXA11* causes dysregulation of calcium homeostasis and stress granule dynamics [16]. Ca2+ homeostasis and SG dynamics are two vital physiological processes that are becoming more and more important in ALS research, and *ANXA11* is a key link between them. C terminal region of *ANXA11* with 4 homologous Conserved Annexin domains, are responsible for phospholipid binding in a calcium-dependent manner and are involved in Ca^2+^ signaling, cell division, apoptosis, and vesicle trafficking [11], [21], *ANXA11* gene mutations can cause abnormal protein aggregation and motor neuron mortality by upsetting the intracellular Ca2+ equilibrium [16]. Both motor neurons and skeletal muscle fibres are susceptible to protein misfolding caused by the *ANXA11* missense mutation [53].

SIGMAR1 is an endoplasmic reticulum protein that regulates variety of molecular functions in various cell types, including the calcium signalling via inositol 1,3,5-triphosphate receptors (IP3 receptors) [26], mitochondrial function, autophagy, ion channel function regulation, lipid transport and prevention of accumulation misfolded proteins [25]. The motor neurons in the brainstem and spinal cord exhibit significant expression of this gene. Several studies have shown an association of several distal neuropathies with severe skeletal muscle dysfuction such as involvement of motor neurons, muscle wasting and atrophy to be linked with mutations in the *SIGMAR1* gene.

Majority of the *In silico* pathogenicity prediction tools like Mutation Taster, Polyphen and SIFT, PROVEAN and SNP&Go predicted these variants to be associated with the disease and highly deleterious in nature. The missense variant of *ANXA11;* p.R452W, has not been associated with ALS till date, to the best of our knowledge. The p.R208W in *SIGMAR1* with a MAF of 0.007 (Global) and 0.019 (South Asian population) respectively and has been previously identified in an old onset Japanese ALS patient [54]. When we analysed and compared the physiochemical properties (hydropathicity, hydrophobicity, molecular weight, polarity, alpha helix, isoelectric point) of the mutated proteins p. R452W and p.R208W respectively vs. the wild type *ANXA11* and *SIGMAR1 respectively*, significant changes were found in both p.R452W and p.R208W variants. Further secondary structure prediction especially in case of mutant p.R452W of *ANXA11* showed significant changes in the confirmation. Superimposition of the wild and mutated type of the protein also revealed that both of the mutants vary considerably from their wild type which is further substantiated by the RMSD and RMSF plots obtained from molecular dynamics simulation results. *In silico* studies predicted that there is significant change in the mutated protein structure compared to its wild type which might result in altered protein function therefore having effect on disease onset and progression. Hence, based on all these results we predicted that the missense variants found in our study are likely pathogenic and have a relationship with the development of disease. Multiple genetic variants may interact among themselves simultaneously to increase risk of ALS, therefore the possibility that both R452W and R208W variant has contributed to the disease pathogenicity can’t be ruled out. Therefore, it is necessary to screen *ANXA11* and *SIGMAR1* in large sample size of young onset ALS patients of Indian origin to determine its frequency in the affected population. Further studies need to be conducted to identify the functional implications of these mutations in ALS patients.

## 5. Conclusion

Our findings broaden the spectrum of ALS causing genes in Indian ALS patients and necessitates neurologists to screen for mutations in *ANXA11* and *SIGMAR1* genes in young onset ALS as well. As ALS is a late onset neurodegenerative disorder where both the genetic mutations accumulating over the years and the environmental factors trigger and predispose the patient towards disease onset, it is important to screen and identify the underlying genetic background of relatively young onset ALS cases where chances of frequency of known ALS candidate gene mutations becomes very high. Moreover, the discovery of a disease-causing gene mutation may eventually result in treatment in the form of personalized medicine in the future.

## Statement and Declarations

### Ethics declaration

This study was performed in line with the principles of the Declaration of Helsinki. Approval was granted by the Ethics Committee of Institute of Science, Banaras Hindu University (*Ref No: I.Sc/ECM-XIV/2022-23/)*

## Acknowledgement

The authors are grateful to the patient and his family members for their contribution to this study. The authors wish to thank Ms. Pooja Sharma, Research Scholar, CSIR-IGIB, New Delhi, India for providing help with the repeat expansion work.

The first author acknowledges Department of Science and Technology (DST), India for providing Junior Research Fellowship (JRF) & Senior Research Fellowship (SRF).

## Author contributions

All authors contributed to the study conception and design. Material preparation, data collection and analysis were performed by Saileyee Roychowdhury, Deepika Joshi, Vinay K Singh, Mohammed Faruq and Parimal Das. The first draft of the manuscript was written by Saileyee Roychowdhury and was checked by Parimal Das. All authors commented on previous versions of the manuscript. All authors read and approved the final manuscript.

## Data Availability

All data generated or analysed during this study are included in this published article.

